# Population-based cervical screening with Human Papillomavirus self-sampling at home and incidence of cervical cancer

**DOI:** 10.64898/2026.01.15.26344201

**Authors:** Helena Andersson, Joakim Dillner

**Affiliations:** Center for cervical cancer elimination, F46, Pathology and Cancer Diagnostics, Medical Diagnostics Karolinska, Karolinska University Hospital and Department of clinical science, intervention and technology, Karolinska Institutet, 141 86 Stockholm, Sweden

**Keywords:** screening, cervical cancer, cancer incidence, self-sampling, Human Papilloma virus, HPV

## Abstract

**Objective:** To describe the Swedish 2020 switch to primary cervical screening using Human Papillomavirus (HPV) self-sampling at home and compare with cervical cancer incidence trends.

**Methods:** Statistics on HPV self-sampling and cervical cancer incidence were obtained from comprehensive nationwide registries.

**Results:** HPV self-sampling was recommended in 2017 for non-attending women and was in 2020 recommended as a primary screening for all women aged 23-70 years of age. During 2016-2020 there were <20,000 annual self-samples taken, but since 2021 >200,000 self-samples are taken annually (about half of all screening tests). During 2015 to 2020 the cervical cancer incidence was stable, between 11 and 12 per 100,000 women. From 2021 and onwards the incidence is declining and was 7,7/100,000 in 2024. The decline is stronger among women younger than 30 (−60%) but is strong in all ages (−27%).

**Conclusions:** A rapid introduction of primary HPV self-sampling at home in 2020-2021 was followed by a rapid decline in invasive cervical cancer incidence.

## Introduction

The elimination of cervical cancer as a public health problem, defined as reducing the cervical cancer incidence to below 4 cases per 100,000 women, is a world-wide goal formulated by the WHO in 2020 [1]. A pillar of the elimination strategy is to use primary HPV-test in cervical cancer screening, with self-sampling being a recommended screening modality [2].

In 2014 WHO recommended HPV as the primary choice for cervical screening [3]. In Sweden, primary HPV screening was recommended for women over 30 in 2015 [4]. HPV self-sampling was recommended for non-attending women in 2017 [5]. During the Covid-19 pandemic most screening appointments were cancelled to avoid crowding in the waiting rooms of the clinics. To prevent a full stop in screening the Swedish National Board of Health and Welfare issued a temporary regulation for screening with primary HPV self-sampling in all ages in 2020 [6]. The regulation became permanent in 2022 [4]. The launch of primary HPV self-sampling improved the screening attendance [7] and was highly cost-effective [8]. Self-sampling has been in routine use for detection of genital infections for many decades and systematic meta-analyses find that it also works well for HPV [9]. The risk for cervical cancer after a negative HPV test (so-called interval cancers) is similar for clinician-taken HPV tests and for self-taken HPV tests [10]. As the strategy both increases screening attendance and has similar cancer-preventive effect as clinician-based sampling, its introduction should result in a decline in cervical cancer incidences. Although correlations may not reflect causality, we reasoned that a simple tabulation of the use of HPV self-sampling and the invasive cervical cancer incidences should give a preliminary indication of whether the strategy works or not.

## Methods

Data on cervical cancer incidence were retrieved from the Swedish Cancer Registry [11]. The registry reports cancer incidences based on the international standard recommended by International Agency for Research on Cancer (IARC) [12].

Data on HPV-tests and HPV self-samples were retrieved from the Swedish National Cervical Screening Registry annual reports at www.nkcx.se. This study is based on already collected information from the Swedish National Cervical Screening Registry. The screening registry gathers data for the purpose of measuring the quality of care, in this case the prevention offered by the cervical cancer screening program. The work conducted by the quality registry is classified as quality assurance in healthcare and does not fall under the Ethics Review Act.

## Results

The number of self-samples and the total number of HPV tests performed within the screening programme during the last decade are reported in Table 1. There is a clear increase in the percentage of HPV tests that are taken as self-samples, from 8% to 49% between 2020 and 2021 (Self-samples were recommended for primary screening for all women in the screening ages in 2020). During 2021-2024 the use of self-samples was stable, at about half of all HPV screening samples. There were >10,000 self-samples taken in 2016, because of a randomized health services study where long-term non-attenders were offered self-samples [13].

**Table 1.**
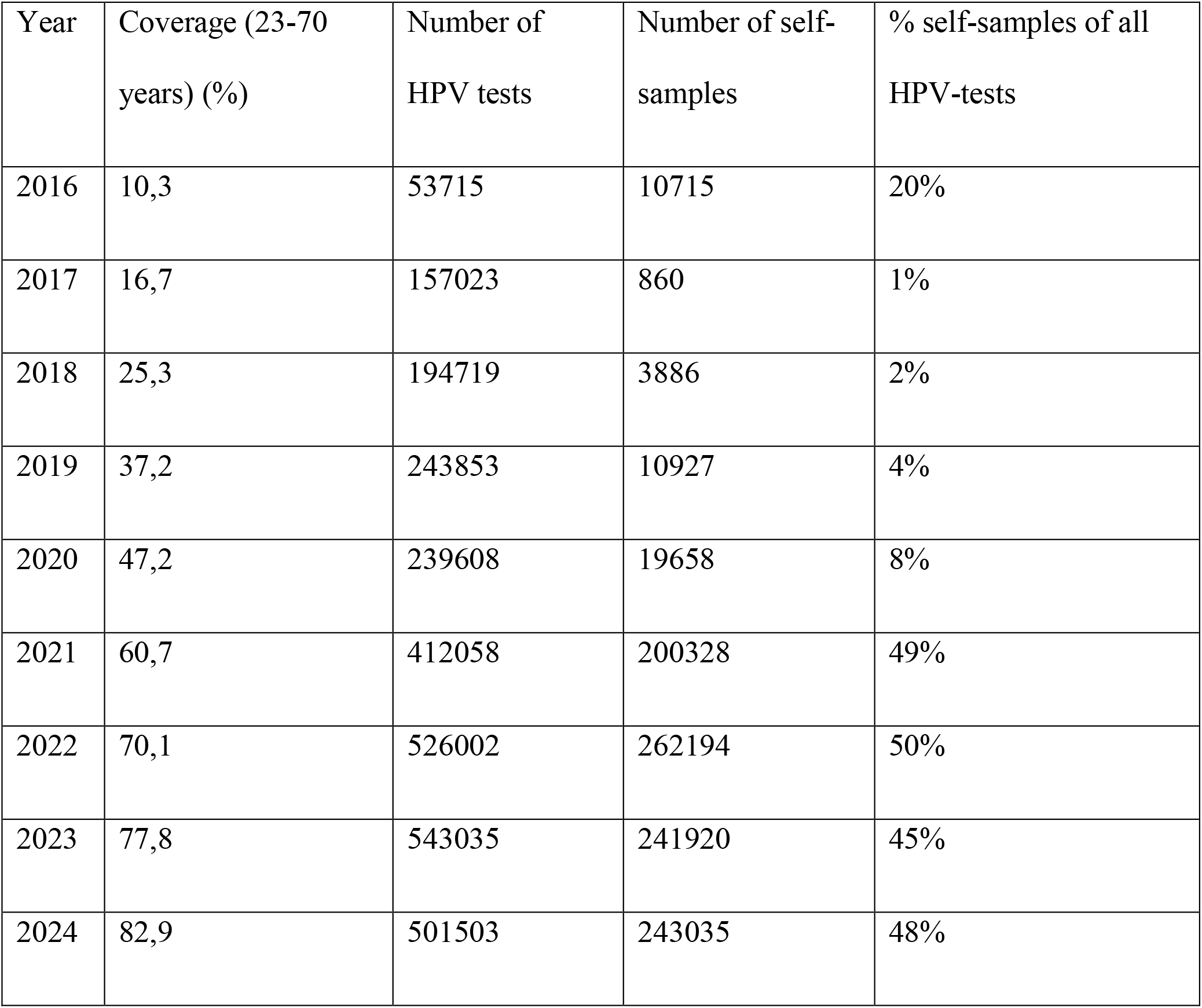
Number of HPV tests taken in Sweden, by calendar year and sampling modality, and in comparison to the total HPV test coverage in the population.

| Year | Coverage (23-70 years) (%) | Number of HPV tests | Number of self-samples | % self-samples of all HPV-tests |
| --- | --- | --- | --- | --- |
| 2016 | 10,3 | 53715 | 10715 | 20% |
| 2017 | 16,7 | 157023 | 860 | 1% |
| 2018 | 25,3 | 194719 | 3886 | 2% |
| 2019 | 37,2 | 243853 | 10927 | 4% |
| 2020 | 47,2 | 239608 | 19658 | 8% |
| 2021 | 60,7 | 412058 | 200328 | 49% |
| 2022 | 70,1 | 526002 | 262194 | 50% |
| 2023 | 77,8 | 543035 | 241920 | 45% |
| 2024 | 82,9 | 501503 | 243035 | 48% |

Between 2015-2020, the overall cervical cancer incidence was stable at 11-12/100,000, but from 2021 to 2024 there was a rapid decline in incidence dropping by more than 27% during this period (Table 2). In the age group 0-29-year-olds there was an almost 60% drop to an incidence of 0,8 in 2024 (Table 2).

**Table 2.** Cervical cancer incidence in Sweden (per 100,000 women) during the period 2015-2024.

| **Year** | **0-29 years old** | **30-85+ years old** | **0-85+ years old** |
| --- | --- | --- | --- |
| **2015** | 3,24 | 16,47 | 11,80 |
| **2016** | 3,03 | 16,09 | 11,47 |
| **2017** | 3,95 | 15,67 | 11,52 |
| **2018** | 2,57 | 15,92 | 11,20 |
| **2019** | 2,45 | 15,82 | 11,12 |
| **2020** | 2,06 | 16,19 | 11,26 |
| **2021** | 1,96 | 15,21 | 10,63 |
| **2022** | 1,79 | 14,72 | 10,29 |
| **2023** | 1,24 | 13,33 | 9,23 |
| **2024** | 0,80 | 11,23 | 7,73 |

## Discussion

We report that the recommendation to use self-sampling for HPV at home in 2020 was rapidly implemented. HPV sampling by health-care personnel is considered an equal alternative to self-sampling and ever since self-sampling was allowed for all women, about half of the screening samples are still taken by healthcare personnel.

We also report a strong decline in cervical cancer incidences during 2021-2024. The decline does correlate with the switch to self-sampling, but there are several additional improvements that may have contributed to the decline. The most important is HPV vaccination, which has greatly reduced prevalences of HPV among women under 30 [14]. In addition, an HPV catch-up vaccination program offering concomitant HPV vaccination and HPV screening to women in the ages 23-31 has been enrolling during 2021-2025 [15]. This, together with the switch to self-sampling has more than doubled the population coverage of HPV tests among women below 30 years of age during 2021-2024 [16]. The incidence trends among women below 30 years of age are therefore likely to reflect both HPV vaccination and HPV self-sampling. Indeed, the decline of the incidences was much larger among the women <30 years of age. There appears to have been some decline among young women already during 2018-2020, which could only be explained by HPV vaccination. The faster decline 2021-2024 among young women could conceivably be attributable to both HPV vaccination and HPV self-sampling.

Among women above 30 years of age, vaccination is likely to have only a limited impact, and the decline is still strong. However, apart from the switch to self-sampling, there are other major improvements ongoing. Notably, cytology is not recommended for any age group since 2022 (no longer considered a screening test). The population coverages of HPV tests (all sampling modalities) have been increasing greatly during the 2016-2024 [16]. It is not straightforward to disentangle how much of this increased HPV testing coverage that is due to the different sampling modalities, but self-sampling appears to be a major contributor [7]. The 2017 recommendation to target non-attending women with self-sampling had a slow start, but has gradually increased in intensity [17, 18] and could also be a contributing cause of the declining incidence trend. Triaging of women positive for high-risk HPV with cytology has been found to be unsafe [2] and has been abandoned, but not until 2025 so this improvement would not have affected the cancer incidences yet.

Systematic reviews find that self-sampling can double the participation in screening compared to clinician-taken samples [17]. As the effect of screening on cancer incidence should be essentially immediate, a population-level switch to self-sampling should result in an immediate reduction in invasive cervical cancer incidence.

## Conclusion

The recommended switch to HPV self-sampling in cervical screening was rapidly implemented and was followed by the expected steep decline in cervical cancer incidences. Although alternative explanations are possible, the reduction in cancer incidences seen suggest that the self-sampling strategy works.

## Funding

This work was supported by the Swedish Cancer Society [grant number 23 2915 Pj]; the Swedish Research Council [grant number 2021-01228_3]; and CIMED [grant number 976207]. The sponsors had no role in the study design, collection, analysis and interpretation of data, writing of the report and decision to submit the article for publication.

## Declaration of competing interest

The authors declare that they have no known competing financial interests or personal relationships that could have appeared to influence the work reported in this paper.

## Data availability

The data used in this study are freely available at NKCx and [Statistikdatabaser - Cancerstatistik - Val](https://sdb.socialstyrelsen.se/if_can/val.aspx)

## Notes

### Competing Interest Statement

The authors have declared no competing interest.

### Funding Statement

This study was funded by the Swedish Cancer Society [grant number 23 2915 Pj]; the Swedish Research Council [grant number 2021-01228_3]; and CIMED [grant number 976207]. The sponsors had no role in the study design, collection, analysis and interpretation of data, writing of the report and decision to submit the article for publication.

### Author Declarations

The study used ONLY openly available human data that were originally located at:https://nkcx.se/index_e.htm and https://sdb.socialstyrelsen.se/if_can/val.aspx

